# Children at familial high risk for schizophrenia or bipolar disorder show sex-specific differences in subcortical grey matter volumes

**DOI:** 10.64898/2026.09.07.26362368

**Authors:** Mathilde Marie Hansen, William F.C. Baaré, Enedino Hernández-Torres, Kit Melissa Larsen, Vasileios Ioakeimidis, Line Korsgaard Johnsen, Nicoline Hemager, Maja Gregersen, Mette Falkenberg Krantz, Nanna Weye, Anne Søndergaard, Christina Bruun Knudsen, Lotte Veddum, Anna Krogh Andreassen, Torben E. Lund, Ole Mors, Anne Amalie Elgaard Thorup, Leif Østergaard, Merete Nordentoft, Hartwig Roman Siebner, Kathrine Skak Madsen

## Abstract

**Background:** Children of parents with schizophrenia (SZ) or bipolar disorder (BP) are at familial high risk (FHR) of developing severe mental illnesses. Based on findings in FHR populations and adults with SZ and BP, we hypothesized that children at FHR for schizophrenia (FHR-SZ) or bipolar disorder (FHR-BP) would show sex-specific differences in subcortical grey matter volume, particularly in the thalamus, amygdala, and hippocampus, compared with population-based controls (PBC).

**Methods:** We examined eight major subcortical grey matter structures in 108 FHR-SZ, 69 FHR-BP, and 120 PBC children aged 11-12 years from the Danish High Risk and Resilience Study. Structural magnetic resonance imaging (MRI) was used to estimate subcortical volumes and R1, a measure of myelin, iron, water, and macromolecular content. Sex-specific group differences were tested across regions.

**Results:** The thalamus and amygdala, but not hippocampus, showed sex-specific group differences in volume. FHR-SZ males showed smaller thalamic volumes than FHR-BP and PBC males, whereas FHR-BP females showed larger thalamic volumes compared with PBC females. FHR-BP males showed larger relative amygdala volumes (corrected for total brain volume) compared to PBC males. No significant differences were observed for subcortical R1.

**Conclusions:** Children at FHR-SZ and FHR-BP exhibited sex-specific differences in subcortical brain volumes before the typical onset of SZ and BP. These differences may reflect early sex-specific neurodevelopmental correlates of familial risk and highlight the importance of modelling sex-specific effects in developmental neuropsychiatric research. Longitudinal studies are needed to determine how these volumetric differences evolve and whether they are associated with later clinical outcomes.

## Introduction

Schizophrenia (SZ) and bipolar disorder (BP) are among the most severe mental disorders, each affecting approximately 1% of the population worldwide.^1,2^ Both disorders are highly heritable and share substantial genetic overlap.^3,4^ A first-degree family history is one of the strongest known risk factors for developing the same or another severe mental illness,^5,6^ with offspring of affected individuals having an estimated 10-40% risk of developing a psychotic or bipolar disorder.^5,7^ Given that SZ and BP are increasingly conceptualized as disorders with specific premorbid neurodevelopmental impairments,^8–10^ studying children at familial high risk (FHR) offers a unique opportunity to investigate the early neurodevelopmental alterations that precede the onset of severe mental disorders.

Neuroimaging studies have identified morphological differences in several subcortical grey matter structures in individuals with SZ and BP.^11,12^ These structures, including the amygdala, nucleus accumbens, hippocampus, and thalamus, play important roles in emotional, executive, and cognitive processes frequently affected in both disorders.^13–18^ Meta-analyses have reported smaller amygdala, nucleus accumbens, hippocampal, and thalamic volumes in adults with SZ compared with healthy controls, with and without adjustment for intracranial volume (ICV).^12,19,20^ Similar, but less pronounced findings have been reported in adults with BP.^11,21^ Findings for other subcortical structures have been less consistent.^12,19,20^ Beyond volumetric alterations, recent quantitative MRI studies have reported altered subcortical tissue properties related to myelin and iron content in adults with SZ,^22^ suggesting that microstructural differences may accompany volumetric abnormalities.

Studies in FHR-SZ and FHR-BP offspring have also reported subcortical brain volumetric differences. A meta-analysis reported smaller volumes of the amygdala, caudate, hippocampus, pallidum, and thalamus in FHR-SZ offspring aged 10-29 years, although these differences were no longer evident after adjustment for ICV.^23^ In FHR-BP offspring aged 11-27 years, the same meta-analysis found smaller relative hippocampal volumes after adjustment for ICV, whereas no differences were observed without adjustment.^23^ Smaller studies of FHR-SZ and FHR-BP offspring have reported heterogeneous findings, including volumetric differences in the amygdala^28,32^ and hippocampus^27,28,31^, as well as null findings across subcortical structures.^24–27^ While informative, these studies generally included relatively small sample sizes and/or broad age ranges spanning childhood to adulthood and did not examine sex differences.^24–32^ Such age ranges encompass periods both before and after the typical age of onset of SZ and BP, as well as different stages of brain maturation during which subcortical structures undergo nonlinear developmental changes.^33,34^ Consequently, it remains unclear whether subcortical volumetric differences are present before the typical age of onset of SZ or BP.

Sex is an important source of heterogeneity in both brain development and severe mental illness. Males and females show different maturational trajectories of subcortical brain structures throughout childhood and adolescence^34,35^ and differ in several clinical characteristics of SZ and BP, including age of onset, symptom presentation, and illness course.^36–39^ Despite this, sex-specific effects have rarely been examined in neuroimaging studies of FHR-SZ and FHR-BP offspring. Recently, we reported sex-specific group differences in brain and cortical volumes in the current sample of 11-12-year-old children, with smaller volumes in FHR-SZ males and larger volumes in FHR-BP females compared with population-based controls (PBC).^40^ Whether similar sex-specific group differences are present in subcortical brain structures remains unknown.

To address these gaps, we investigated subcortical grey matter volumes in 11-12-year-old children in the prospective Danish High Risk and Resilience Study (VIA),^41^ a large cohort of children born to parents with SZ, BP, or PBC. Given our previous finding of sex-specific brain volume differences in the same cohort,^40^ established sex differences in subcortical brain development,^34,35^ and sex differences in the clinical presentation of SZ and BP,^36–39^ we hypothesized that group differences in subcortical volumes would be sex-specific. We therefore first tested group-by-sex interactions, followed by group differences. Based on findings in adults with SZ or BP and previous FHR studies, we focused a priori on the amygdala, hippocampus, and thalamus. The nucleus accumbens, caudate, pallidum, and putamen were examined exploratorily. We additionally examined whether group differences were associated with lifetime Axis-I diagnosis. Finally, we explored group differences in the subcortical longitudinal relaxation rate (R1), reflecting tissue composition, including myelin, iron, water, and macromolecular content.^42^ Given the sparse literature in FHR populations, no specific hypotheses were formulated for these analyses.

## Methods

### Participants

This study is a part of the Danish High Risk and Resilience Study - VIA, a prospective, nationwide cohort study investigating developmental pathways of psychiatric disorders.^43^ At baseline (VIA 7), the cohort included 522 7-year-old children with at least one parent diagnosed with SZ (FHR-SZ, n=202), BP (FHR-BP, n=120) or neither disorder (referred to as PBC, n=200). SZ was defined according to ICD-10 codes F20, F22, and F25 or ICD-8 codes 295, 297, 298.29, 298.39, 298.89, and 298.99. BP was defined as ICD-10 codes F30 and F31 or ICD-8 codes 296.19 and 296.39. The families were reinvited to participate in the second wave of assessment (VIA 11) when the children were 11 years old, with 465 children participating (FHR-SZ n=179, FHR-BP n=105, PBC n=181), corresponding to a retention rate of 89.1 %. MRI was added to the assessment battery at VIA 11. The present study included structural MRI data from 297 children from the VIA 11 study (FHR-SZ, n=108; FHR-BP, n=69; PBC, n=120). The flowchart in Figure S1 summarizes the inclusion and exclusion procedures. Additional details are provided in the VIA study protocols,^41,43^ and VIA recruitment process paper.^44^ Written informed consent was obtained from all parents or legal guardians prior to participation. The VIA 11 study protocol was approved by the Danish National Committee on Health Research Ethics (Protocol H16043682) and the Danish Data Protection Agency (ID: RHP-2017-003, I-suite: 05333) and conducted in accordance with the Declaration of Helsinki.

### Clinical characteristics

Participants underwent a comprehensive clinical and behavioral evaluation.^45^ The Child Behavior Checklist (CBCL), school-age version, completed by caregivers, assessed the children’s behavioral and emotional problems.^46^ The Children’s Global Assessment Scale (CGAS) evaluated global functioning during the preceding month,^47^ based on caregiver-child interviews. Lifetime Axis-I diagnoses (excluding elimination disorders, transient/unspecified tics, and specific phobias) were assessed using the semi-structured face-to-face interview the Kiddie Schedule for Affective Disorders and Schizophrenia for School-Age Children - Present and Lifetime Version (K-SADS-PL),^48^ followed by a clinical conference with a child and adolescent psychiatrist. Within the FHR-SZ group, three children met criteria for an Axis-I psychotic disorder (DSM 298.9; none with 298.8/297.1/292.30/295.90). No cases of mania (DSM 296) were identified. Pubertal stage was assessed using self-reported Tanner staging after brief instructions, based on the highest ratings across genital/breast and pubic hair.^49^ Additionally, we used the Personal and Social Performance (PSP) Scale interview to assess the primary caregivers’ level of functioning.^50^

### MRI protocol

Participants underwent structural and functional whole-brain MRI protocol on 3T Siemens scanners at one of two sites: the Danish Research Centre for Magnetic Resonance (DRCMR) in Copenhagen or the Centre of Functionally Integrative Neuroscience (CFIN) in Aarhus. Each scanning session lasted about 75 minutes. High-resolution T1-weighted images were acquired using 3D magnetization prepared 2 rapid acquisition gradient echoes (MP2RAGE) with incorporated fat image navigators to enable retrospective motion correction,^51^ with harmonized sequence parameters across sites (see Supplementary Methods). Raw images were visually inspected by trained personnel, blinded to group membership. Signs of brain pathology were reviewed with a neuroradiologist and referred to medical auspices if relevant. We excluded one child due to incidental findings and 16 children due to poor image quality (Figure S1).

### Segmentation of subcortical structures

The MP2RAGE images were processed using the approach of J.P. Marques^52^ to generate T1-weighted images for segmentation and R1 (1/T1) maps. The segmentation of the eight subcortical structures was performed with three different tools, selected for their reported performance in segmenting these structures.^53–55^ Sequence Adaptive Multimodal Segmentation (SAMSEG)^56^ and the hypothalamic subunits segmentation,^57^ both implemented in FreeSurfer 7.2, were used to segment the bilateral thalamus, caudate, nucleus accumbens, hippocampus, amygdala, and hypothalamus. FIRST^58^ in FSL, was used to segment the bilateral putamen and pallidum. Volumes of all eight subcortical structures were extracted, and bilateral averages were used in our statistical analyses. Total brain volume (TBV) was segmented with SAMSEG and extracted to adjust for overall brain size. Finally, R1 averages were calculated for each bilateral subcortical structure for each participant. One participant was excluded due to incomplete R1 data, leaving 296 children for the R1 analysis.

### Quality control of the subcortical segmentations and final dataset

Visual quality control was performed using an in-house graphical user interface, with group status blinded. Poorly segmented structures were excluded, and three participants with more than four poor-quality labels were excluded from further analyses. The hippocampus was inadequately segmented in 124 children, with volumes overestimated due to non-systematic inclusion of the cortex within the parahippocampal sulcus. These cases were manually edited in one or both hippocampal labels using FSLeyes (FSL version 6.0.4). The Total Euler Number (TEN) was extracted with FreeSurfer and used as a proxy for image quality.^59^ Three scans with a TEN more than three standard deviations below the mean were excluded.

The cohort with valid structural MP2RAGE scans included five sibling pairs. To ensure independence, the most recently enrolled sibling from each pair was excluded, yielding a final sample of 297 children (Figure S1). Due to structure-specific exclusions, the number of available data for each bilateral subcortical structure ranged from 269 to 296 (Table 2 and 3). Exclusions did not significantly differ between groups (Table S1).

### Statistical analysis

Analyses were conducted in R, version 4.0.4,^60^ primarily using the *car*^61^ and *lsmeans*^62^ packages. Demographic and clinical characteristics were compared across groups using chi-square tests and one-way analyses of variance, followed by pairwise comparisons. Drop-out analyses employed t-tests and chi-square tests.

To test our hypothesis, we assessed group-by-sex and group effects on bilateral subcortical volumes using two Analysis of Covariance (ANCOVA) models. In Model 1, we tested the group-by-sex interaction and adjusted for age at MRI, site, and TEN (*volume ∼ group x sex + group + sex + age + site + TEN*). If the interaction was not significant, we removed the interaction term to test the main effect of group (Model 2), adjusting for age, sex, site, and TEN (*volume ∼ group + age + site + TEN*). Pairwise group comparisons (FHR-SZ vs. PBC; FHR-BP vs. PBC; FHR-BP vs. FHR-SZ) were conducted separately in males and females, using t-tests on the estimated marginal means from *lsmeans*. We first examined our *a priori* regions-of-interest (thalamus, hippocampus, and amygdala). We then explored group differences in the remaining subcortical volumes (nucleus accumbens, caudate, hypothalamus, pallidum, and putamen). The Benjamini-Hochberg false discovery rate (FDR) procedure^63^ was applied to adjust for multiple comparisons (q*≤*0.05). We first corrected for three *a priori* volumes and subsequently for eight volumes (i.e., three *a priori* and five additional structures). Finally, analyses were repeated to examine relative volume differences by including total brain volume (TBV) as a covariate, following recommended practices for pediatric neuroimaging.^64^ Significant findings from the primary analysis were followed up with additional analyses, using an uncorrected p*≤*0.05. First, we evaluated whether significant group-by-sex or group effects persisted after controlling for pubertal stage. Second, we examined whether group differences were associated with lifetime Axis-I diagnosis. To do this, we first included lifetime Axis-I diagnosis as a covariate in the ANCOVA model. Next, we stratified FHR-SZ, FHR-BP, and PBC groups into subgroups *with* (+) or *without* (÷) an Axis-I diagnosis and conducted pairwise t-tests. Third, we examined the unilateral volumes of the structures showing significant bilateral differences to determine whether the effects were mainly driven by one hemisphere. Finally, we conducted exploratory analyses of bilateral R1 values using the same approach.

Effect sizes were quantified using partial eta², which indicates how much of the variability in the dependent variable is explained by the predictor. We interpreted eta²-values as negligible (<0.01), small (0.01-0.06), medium (0.06-0.14), and large (≥0.14) effect sizes, according to convention.^65^ For pairwise comparisons, Cohen’s *d* were calculated and interpreted as small (0.2), medium (0.5), and large (0.8) effect sizes.^66^ In addition, we complemented our analyses with a Bayesian framework by calculating Bayes factors (BF_10_), quantifying evidence for the alternative hypothesis (H₁) relative to the null hypothesis (H₀). A BF_10_ of 1 indicates no evidence for either hypothesis. Increasing values indicate anecdotal (>1-3), moderate (>3-10), strong (>10-30), very strong (>30-100), and decisive (>100) evidence for H₁, while decreasing values below 1 indicate increasing evidence for H₀.^67,68^

## Results

### Demographic and clinical variables

Demographic and clinical characteristics are summarized in Table 1. The FHR-SZ, FHR-BP, and PBC groups did not significantly differ in age, sex, or MRI scan site. The FHR groups exhibited higher CBCL scores, indicating more behavioral and emotional problems, and lower CGAS scores, indicating poorer global functioning, compared to the PBC group. The FHR groups also had a higher prevalence of lifetime Axis-I diagnoses than the PBC group, consistent with previous VIA 11 reports.^45,69^

**Table 1.**
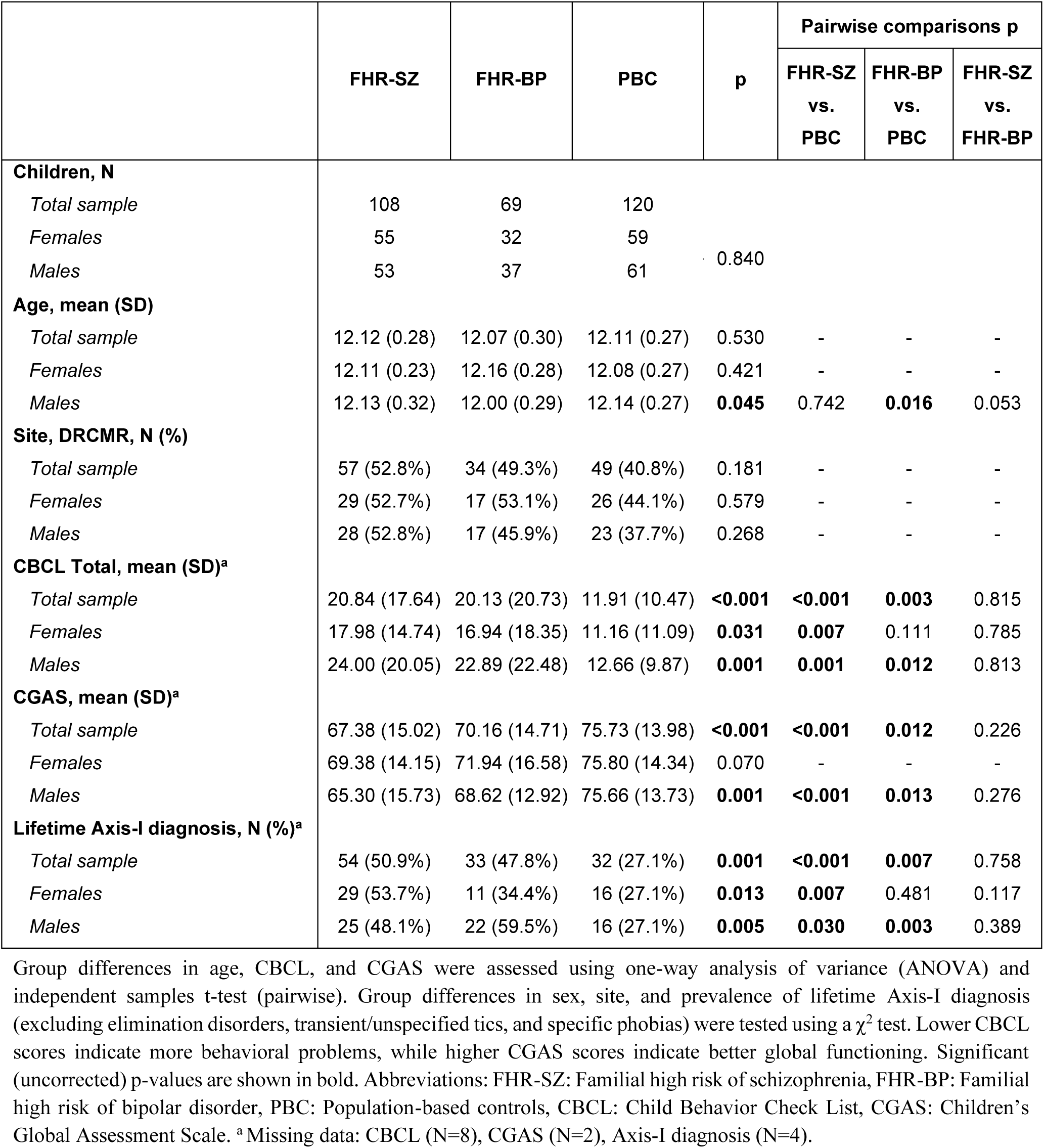
Demographic and clinical characteristics and group differences between the FHR-SZ, FHR-BP, and PBC groups for the total sample, females, and males.

**Table 2.** Results from the *a priori* hypothesis of group-by-sex and group effects in the amygdala, hippocampus, and thalamus.

|  |  | Group |  |  |  |  | Group*sex |  |  |  |  |
| --- | --- | --- | --- | --- | --- | --- | --- | --- | --- | --- | --- |
| Structure (N) | Model | *Eta*2 | *F* | *p* | *q* | *BF*10 | *Eta*2 | *F* | *p* | *q* | *BF*10 |
| Without TBV |  |  |  |  |  |  |  |  |  |  |  |
| Amygdala (290) | 1 | 0.000 | 0.059 | 0.942 | - | - | 0.032 | 2.681 | 0.070 | - | 0.671 |
|  | 2 | 0.017 | 2.451 | 0.088 | 0.088 | 0.351 | - | - | - | - | - |
| Hippocampus (291) | 1 | 0.000 | 0.012 | 0.988 | - | - | 0.036 | 3.014 | 0.051 | - | 0.929 |
|  | 2 | 0.021 | 3.041 | 0.049 | 0.074 | 0.627 | - | - | - | - | - |
| Thalamus (296) | 1 | 0.020 | 2.927 | 0.055 | - | - | 0.031 | 4.653 | 0.010 | **0.030** | 4.400 |
| With TBV |  |  |  |  |  |  |  |  |  |  |  |
| Amygdala (290) | 1 | 0.009 | 1.296 | 0.275 | - | - | 0.030 | 4.338 | 0.014 | **0.042** | 2.373 |
| Hippocampus (291) | 1 | 0.007 | 0.973 | 0.379 | - | - | 0.008 | 1.115 | 0.329 | - | 0.173 |
|  | 2 | 0.006 | 0.798 | 0.451 | 0.451 | 0.082 | - | - | - | - | - |
| Thalamus (296) | 1 | 0.010 | 1.416 | 0.244 | - | - | 0.007 | 0.329 | 0.720 | - | 0.095 |
|  | 2 | 0.015 | 2.242 | 0.108 | 0.162 | 0.284 | - | - | - | - | - |

Subcortical grey matter structures *without* and *with* adjustment for total brain volume (TBV). Model 1: ANCOVA with group-by-sex, group, sex, age, site, and Total Euler Number. Model 2: ANCOVA with group, sex, age, site, and Total Euler Number. FDR-corrected q-values are reported for the group-by-sex model (Model 1) when $p < 0.05$ ; otherwise, they are reported for the group model (Model 2). Significant q values are shown in bold. Abbreviations: $BF_{10}$ : Bayes Factor.

**Table 3.**
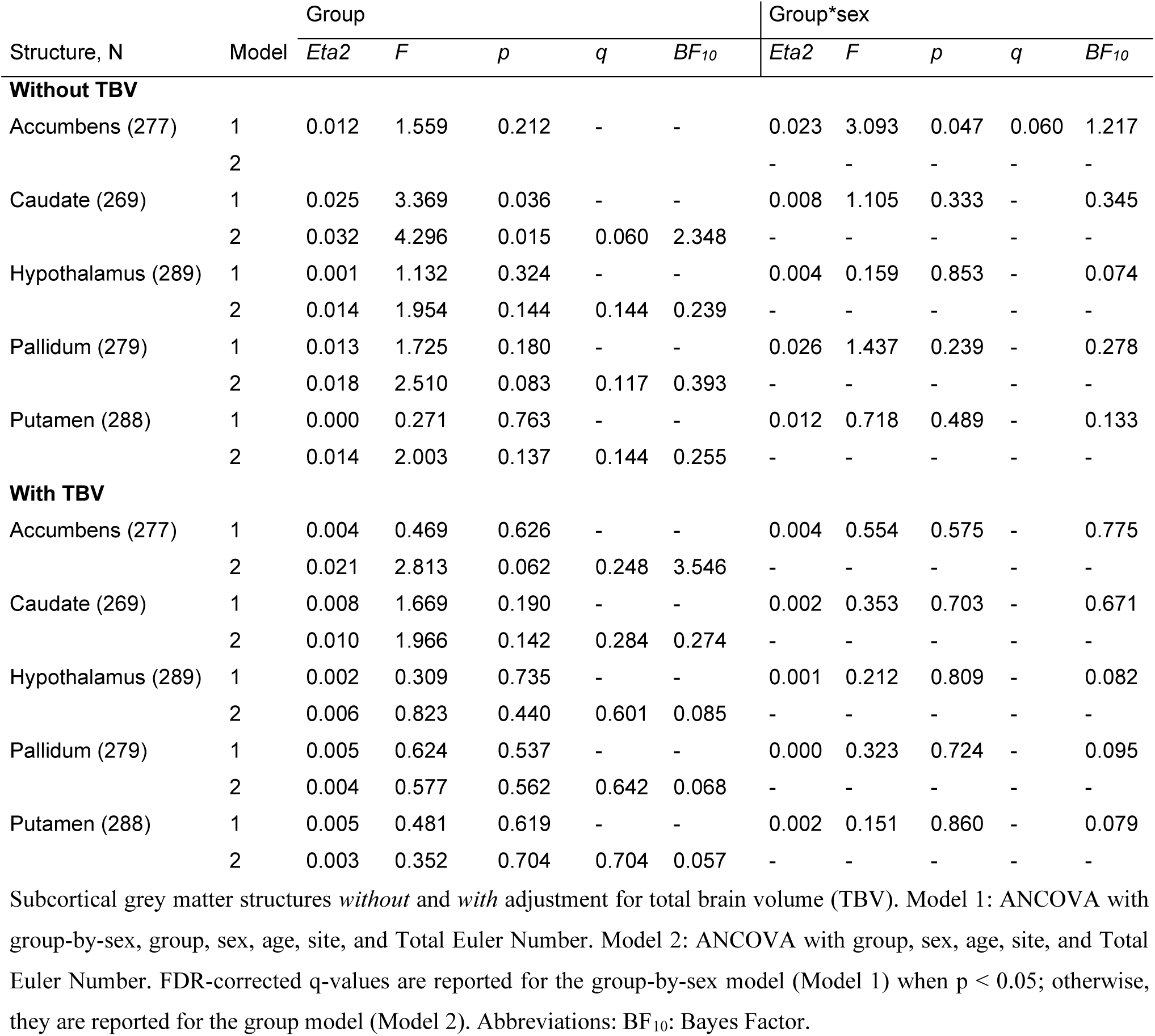
Results from the exploratory analysis of group-by-sex and group effects in the accumbens, caudate, hypothalamus, pallidum, and putamen.

| Structure, N | Model | Group |  |  |  |  | Group*sex |  |  |  |  |
| --- | --- | --- | --- | --- | --- | --- | --- | --- | --- | --- | --- |
|  |  | *Eta*2 | *F* | *p* | *q* | *BF*10 | *Eta*2 | *F* | *p* | *q* | *BF*10 |
| Without TBV |  |  |  |  |  |  |  |  |  |  |  |
| Accumbens (277) | 1 | 0.012 | 1.559 | 0.212 | - | - | 0.023 | 3.093 | 0.047 | 0.060 | 1.217 |
|  | 2 |  |  |  |  |  | - | - | - | - | - |
| Caudate (269) | 1 | 0.025 | 3.369 | 0.036 | - | - | 0.008 | 1.105 | 0.333 | - | 0.345 |
|  | 2 | 0.032 | 4.296 | 0.015 | 0.060 | 2.348 | - | - | - | - | - |
| Hypothalamus (289) | 1 | 0.001 | 1.132 | 0.324 | - | - | 0.004 | 0.159 | 0.853 | - | 0.074 |
|  | 2 | 0.014 | 1.954 | 0.144 | 0.144 | 0.239 | - | - | - | - | - |
| Pallidum (279) | 1 | 0.013 | 1.725 | 0.180 | - | - | 0.026 | 1.437 | 0.239 | - | 0.278 |
|  | 2 | 0.018 | 2.510 | 0.083 | 0.117 | 0.393 | - | - | - | - | - |
| Putamen (288) | 1 | 0.000 | 0.271 | 0.763 | - | - | 0.012 | 0.718 | 0.489 | - | 0.133 |
|  | 2 | 0.014 | 2.003 | 0.137 | 0.144 | 0.255 | - | - | - | - | - |
| With TBV |  |  |  |  |  |  |  |  |  |  |  |
| Accumbens (277) | 1 | 0.004 | 0.469 | 0.626 | - | - | 0.004 | 0.554 | 0.575 | - | 0.775 |
|  | 2 | 0.021 | 2.813 | 0.062 | 0.248 | 3.546 | - | - | - | - | - |
| Caudate (269) | 1 | 0.008 | 1.669 | 0.190 | - | - | 0.002 | 0.353 | 0.703 | - | 0.671 |
|  | 2 | 0.010 | 1.966 | 0.142 | 0.284 | 0.274 | - | - | - | - | - |
| Hypothalamus (289) | 1 | 0.002 | 0.309 | 0.735 | - | - | 0.001 | 0.212 | 0.809 | - | 0.082 |
|  | 2 | 0.006 | 0.823 | 0.440 | 0.601 | 0.085 | - | - | - | - | - |
| Pallidum (279) | 1 | 0.005 | 0.624 | 0.537 | - | - | 0.000 | 0.323 | 0.724 | - | 0.095 |
|  | 2 | 0.004 | 0.577 | 0.562 | 0.642 | 0.068 | - | - | - | - | - |
| Putamen (288) | 1 | 0.005 | 0.481 | 0.619 | - | - | 0.002 | 0.151 | 0.860 | - | 0.079 |
|  | 2 | 0.003 | 0.352 | 0.704 | 0.704 | 0.057 | - | - | - | - | - |

Subcortical grey matter structures *without* and *with* adjustment for total brain volume (TBV). Model 1: ANCOVA with group-by-sex, group, sex, age, site, and Total Euler Number. Model 2: ANCOVA with group, sex, age, site, and Total Euler Number. FDR-corrected q-values are reported for the group-by-sex model (Model 1) when $p < 0.05$ ; otherwise, they are reported for the group model (Model 2). Abbreviations: $BF_{10}$ : Bayes Factor.

Drop-out analyses comparing the present sample to the full VIA 11 cohort showed that the included children at FHR-SZ and FHR-BP had higher CGAS scores than those not included (p<0.047). Moreover, children at FHR-SZ had lower total CBCL scores (p=0.016), and their primary caregiver had higher PSP scores (p=0.013), indicating better functioning among included FHR-SZ children and their primary caregiver (Table S2 and S3).

### A priori hypothesis: Sex-specific group differences in thalamus, amygdala, and hippocampus volumes

Results from the primary models are presented in Table 2. A significant group-by-sex effect was observed for thalamus volume that survived FDR correction and showed moderate Bayesian evidence, but the effect was no longer significant after TBV adjustment. A significant group-by-sex effect was also observed for relative amygdala volume, which survived FDR correction. However, Bayesian evidence was only anecdotal. No significant group-by-sex or group effects were observed for hippocampus volumes.

Adjustment for pubertal stage and the time interval between MRI and pubertal assessment did not alter the group-by-sex effects in thalamus and relative amygdala volumes (p<0.023; Table S4). However, there was substantial variability in the time interval (range: −46.4 to 42.6 weeks; median interval: 3.6 weeks; IQR: 7 weeks).

To clarify the observed group-by-sex interactions, we conducted pairwise comparisons for males and females separately. Males at FHR-SZ had significantly smaller thalamus volumes than both PBC and FHR-BP males, with strong Bayesian evidence (p<0.002, BF_10_>23.8, Figure 1 and S2, Table S5). Females at FHR-BP had significantly larger thalamus volumes than PBC females, but the Bayesian evidence was only anecdotal (p=0.013, BF_10_=3.1, Figures 1 and S2, Table S6). For relative amygdala volume, males at FHR-BP exhibited significantly larger volumes than PBC males with strong Bayesian evidence (p=0.006, BF_10_=11.4, Figures 1 and S2, Table S5), while no significant differences were observed for males at FHR-SZ (Table S5), or for any of the female groups (Table S6). Pairwise comparisons showed that males at FHR-SZ had smaller hippocampal volumes than FHR-BP and PBC males with moderate Bayesian evidence (Figure 1 and S2, Table S5).

**Figure 1.**
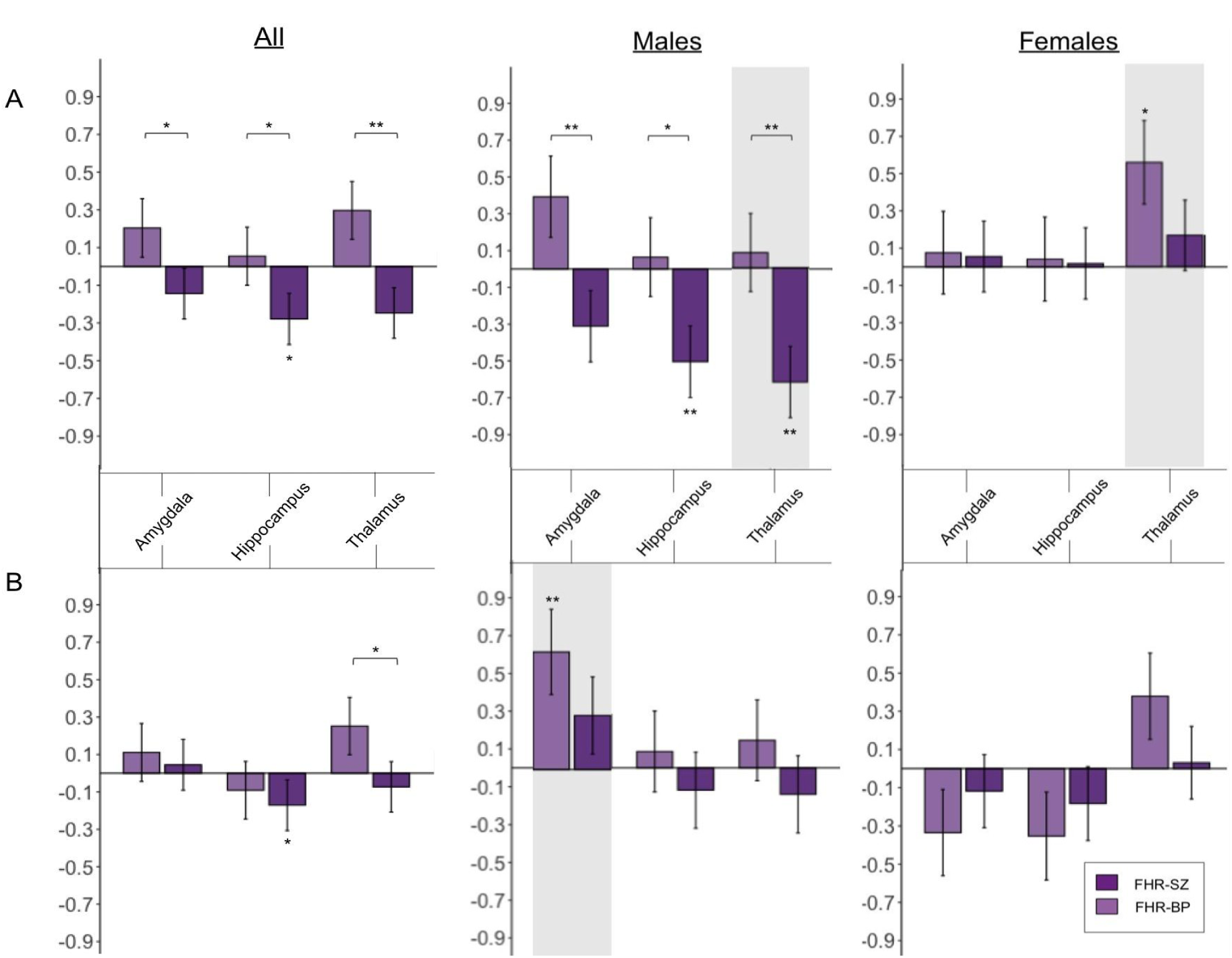
Results of the pairwise analysis of the *a priori* subcortical volumes in the whole group (all, left), males (middle), and females (right). Effect sizes (Cohen’s *d*) are shown for t-test comparing FHR-BP (light purple) or FHR-SZ (dark purple) vs. PBC A) without and B) with adjustment for total brain volume (TBV). Vertical bars denote the standard error of the effect size. P-values are annotated as p<0.05*; p <0.01**, p<0.001***. Grey shading highlights significant FDR-corrected group differences (q< 0.05) for our *a priori* hypothesis. Brackets indicate significant differences between the FHR-SZ and FHR-BP groups. Abbreviations: FHR-SZ: Familial high risk of schizophrenia. FHR-BP: Familial high risk of bipolar disorder. PBC: Population-based controls.

For comparison with previous studies, we also examined whole-sample group differences without the group-by-sex interaction. These largely reflected the sex-specific findings (Figures 1-2, Supplementary Results; Table S7).

**Figure 2.**
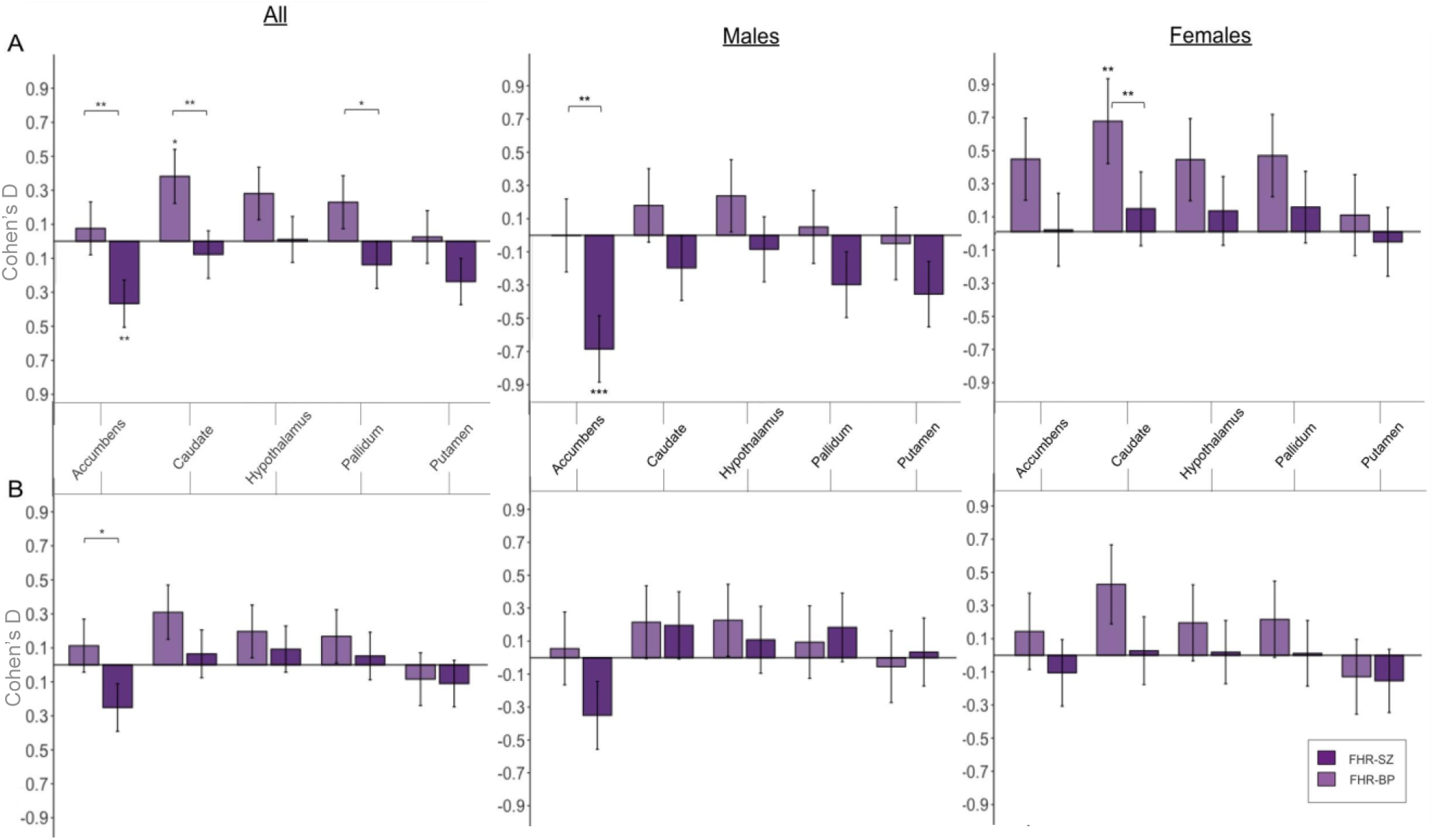
Results of the pairwise analysis of the exploratory subcortical volumes in the whole group (all, left), males (middle), and females (right). Effect sizes (Cohen’s *d*) are shown for the t-test comparing FHR-BP (light purple) or FHR-SZ (dark purple) vs. PBC A) without and B) with adjustment for total brain volume (TBV). P-values are annotated as p<0.05*; p <0.01**, p<0.001***. Brackets indicate significant differences between the FHR-SZ and FHR-BP groups. Abbreviations: FHR-SZ: Familial high risk of schizophrenia. FHR-BP: Familial high risk of bipolar disorder. PBC: Population-based controls.

### Exploratory analysis of accumbens, caudate, hypothalamus, pallidum, and putamen volumes

Results of the exploratory analyses are presented in Figures 2 and S2, and Tables 3 and S5-S7. A group-by-sex effect for nucleus accumbens volume and a group effect for caudate volume were observed, although neither survived FDR correction. Pairwise analysis showed strong Bayesian evidence for smaller nucleus accumbens volumes in FHR-SZ males than in FHR-BP and PBC males (p≤0.003, BF_10_≥21.4), representing one of the largest effect sizes (*d≤*-0.684) observed among the examined subcortical structures. Females at FHR-BP exhibited anecdotal to moderate Bayesian evidence for larger caudate volume than FHR-SZ and PBC females (p≤0.046, BF_10_≥1.5). We found no significant group-by-sex or group differences for the hypothalamus, pallidum, or putamen, and BF_10_ provided no evidence for differences.

### Role of lifetime Axis-I diagnosis in thalamus and amygdala volume differences

Including lifetime Axis-I diagnosis as a covariate did not alter the pattern of results, suggesting that the observed group differences in thalamus and relative amygdala volume were not explained by Axis-I diagnosis (Table S8). Next, children were stratified into subgroups *with* and *without* a lifetime Axis-I diagnosis within each group (Table S9). No differences were observed within the FHR groups (p>0.161). Within the PBC group, males with an Axis-I diagnosis exhibited larger relative amygdala volumes than those without a diagnosis (p=0.015).

### Contribution of the left and right subcortical volumes

To assess lateralization, pairwise analyses were performed separately for left and right subcortical volumes (Table S10). The observed thalamic and relative amygdala volume differences were significant in both hemispheres (p<0.023), mirroring the bilateral findings (Table S10).

### Exploratory subcortical R1 analyses

In exploratory analyses of R1 values, we observed uncorrected group-by-sex effects in the amygdala and hippocampus, and an uncorrected group effect in the nucleus accumbens. However, none survived FDR correction, and the Bayesian evidence was anecdotal. No significant effects were detected in the caudate, pallidum, or hypothalamus. Results are presented in Table S11.

## Discussion

In the largest MRI study to date of 11-12-year-old children at FHR-SZ and FHR-BP, we found sex-specific differences in subcortical grey matter volumes. Males at FHR-SZ exhibited smaller thalamic volumes than FHR-BP and PBC males, while females at FHR-BP showed larger thalamic volumes than PBC females. In addition, males at FHR-BP exhibited larger relative amygdala volumes than PBC males. Exploratory analyses further suggested smaller nucleus accumbens volumes in males at FHR-SZ, with effect sizes comparable to those of the thalamic findings, although these findings should be interpreted cautiously given their exploratory nature. The observed volumetric differences were independent of lifetime Axis-I diagnosis, supporting their potential role as neurodevelopmental markers of familial risk rather than consequences of early psychopathology. No significant differences were detected in the exploratory analysis of region-specific subcortical R1 values.

### Smaller subcortical volumes in males at FHR-SZ

Males at FHR-SZ exhibited smaller thalamic volumes than both FHR-BP and PBC males. Results from exploratory analyses further suggested smaller nucleus accumbens volumes, with modest reductions observed in the hippocampus. Similar to our findings, a meta-analysis of FHR-SZ offspring reported smaller thalamic volumes and nominally smaller hippocampal volumes before adjustment for ICV.^23^ Studies of adults with SZ have likewise reported smaller thalamic, hippocampal, and nucleus accumbens volumes, although without considering potential sex-specific effects.^12,19,20^ Studies of group-by-sex effects in FHR-SZ offspring are lacking, making it difficult to determine whether previously reported group-level differences may have been driven primarily by males. The loss of significance after TBV adjustment suggests that the thalamic and nucleus accumbens differences largely reflect a broader pattern of reduced brain size rather than structure-specific volumetric differences. This interpretation is consistent with our previous findings from the same cohort, where males at FHR-SZ exhibited smaller intracranial, brain, and cortical volumes.^40^ Given that ICV is largely established during early brain development and is closely linked to overall brain growth in childhood,^70–72^ our findings may reflect alterations in early neurodevelopmental processes. Nevertheless, the volume reductions were not uniformly distributed across subcortical structures, with the strongest effects observed in the thalamus and nucleus accumbens, suggesting that some structures may be more affected than others. The absence of similar findings in females suggests that familial risk for SZ may interact with sex-specific developmental processes. Although the mechanisms underlying these sex differences remain unclear, emerging evidence suggests that the phenotypic expression of liability for psychotic disorders may differ between males and females, with polygenic burden showing stronger associations with psychosis risk in males.^73^ Our findings raise the possibility that such sex-dependent effects may also extend to neurodevelopmental brain measures.

### Larger subcortical volumes in FHR-BP

Females at FHR-BP showed larger thalamic volumes than PBC females, while males at FHR-BP exhibited larger relative amygdala volumes (i.e., adjusted for TBV) than PBC males. The larger thalamic volumes parallel our previous findings from the same cohort, where FHR-BP females exhibited larger brain and cortical volumes than PBC females,^40^ as well as studies reporting larger intracranial and brain volumes in first-degree relatives of individuals with BP, although potential sex differences were not examined.^23^ Similarly, a twin study reported a positive association between genetic liability for BP and intracranial volume, suggesting that larger brain volumes may be associated with familial liability for BP.^74^ In contrast, the larger relative amygdala volume in FHR-BP males is particularly noteworthy, as it suggests a region-specific difference rather than a reflection of overall brain size. Consistent with our findings, larger right amygdala volumes have previously been reported in unaffected offspring (6-17 years) of parents with BP compared with both offspring with a psychiatric diagnosis and healthy controls, although sex-specific effects were not examined.^28^ While that study suggested that amygdala enlargement was confined to unaffected offspring, our findings persisted after accounting for lifetime Axis-I diagnosis, suggesting that the observed association was not attributable to early psychopathology but may instead reflect neurodevelopmental correlates of familial risk. Meta-analyses of adults with BP have generally reported smaller amygdala and thalamic volumes than controls.^21,23^ However, a large ENIGMA study identified a sex-by-diagnosis interaction, with larger thalamic volumes in adult females with BP, consistent with our finding in females at FHR-BP.^11^ Together, these findings suggest that the subcortical volumetric correlates of familial liability may differ across brain regions, with some findings, such as larger thalamic volumes in females, also being observed in adulthood, while others, including the amygdala, differ from the pattern typically reported in adult BP. Longitudinal follow-up will be essential to determine how these volumetric differences evolve across adolescence and whether they are associated with later psychopathology.

### Strengths and Limitations

A key strength of this study is the VIA cohort, comprising a large, well-characterized sample of same-aged children identified through national registers, enhancing representativeness. The narrow age range reduces developmental heterogeneity and enables investigation of neuroanatomical differences before the typical age of onset of severe mental illness, thereby minimizing potential confounding effects of chronic illness and prolonged psychotropic medication use. The inclusion of both FHR-SZ and FHR-BP groups, together with a population-based comparison group containing a proportion of children with Axis-I diagnoses, enhances representativeness, and reduces the likelihood of an artificially healthy control group. Furthermore, extensive clinical characterization enabled follow-up analyses demonstrating that the observed volumetric differences persisted after accounting for lifetime Axis-I disorders. Data processing adhered to current recommendations regarding quality control,^59^ computational segmentation tools,^53,54^ and statistical analyses, and Bayesian analyses were used alongside frequentist statistics to provide complementary evidence. Although post hoc harmonization methods were not applied, the use of harmonized MRI protocols, centralized preprocessing and quality control, and balanced group distributions across sites likely reduced site-related variability without introducing a risk of overcorrection.

Several limitations should be considered. The smaller FHR-BP sample may limit power to detect subtle effects, particularly in sex-stratified analyses. Participant heterogeneity and limited power for disorder-specific analyses led to the use of broad Axis-I categorizations, which may obscure disorder-specific patterns. Attrition analyses indicated that the included FHR-SZ children and their primary caregivers exhibited better functioning than those not included, suggesting potential selection bias and possible underestimation of group differences. Although FHR children are at increased risk of broad psychopathology, most will not develop SZ or BP, and the present findings should therefore not be interpreted as markers of specific future illness but rather as neurostructural indicators of possible mental vulnerability. In addition, the temporal gap between MRI and pubertal assessment, together with the use of self-reported Tanner staging, may have reduced the precision of pubertal measures. Finally, the cross-sectional design precludes conclusions regarding developmental trajectories and the predictive significance of the observed volumetric differences.

## Conclusions

This study identified distinct sex-specific patterns of subcortical volumetric differences in 11–12-year-old children at FHR-SZ and FHR-BP. Males at FHR-SZ exhibited smaller thalamic volumes, whereas females and males at FHR-BP showed larger thalamic and relative amygdala volumes, respectively. These findings support the presence of sex-specific brain differences associated with familial risk before the typical age of onset of SZ and BP. Longitudinal, and sex-specific, follow-up is needed to determine how these differences evolve across adolescence and whether they are associated with later clinical outcomes.

## Data Availability

Due to Danish legislation, we cannot offer access to data unless there is a specific collaboration agreement with the research group.

## Acknowledgements

We would like to thank the children and their families for their participation in the Danish High Risk and Resilience Study. We are grateful to Simon Yamazaki Jensen for support with statistical analyses, Line Carmichael for coordinating participant recruitment, and Jessica Ohland for managing the study database. We also acknowledge Agnete Albertsen, Anna Møller, Benthe Vink, Daban Sulaiman, Gøkze Akkas, Jonas Ingerslev, Malte Lundby, and Natascha Larsen at the DRCMR, as well as Anette Bundgaard, Henriette Stadsgaard, Merete Birk, Nanna Steffensen, and Oskar Jefsen at CFIN, for their valuable contributions to MRI data collection. We thank Daniel Gallichan for providing access to the FatNavs-MP2RAGE sequence. Language editing support was provided by OpenAI’s ChatGPT (v5), which assisted with refinement of sentence structure and grammar to improve clarity. This support was limited to text refinement and did not influence the scientific content or conclusions of the study.

## Funding/Support

This study was supported by the Independent Research Fund Denmark (3166-00143B), the Research Fund in the Capital Region of Denmark, the Lundbeck Foundation (R155-2014-1724), Innovation Fund Denmark (6152-00002B), Beatrice Surovell Haskell Fund for Child Mental Health Research of Copenhagen (11531), the Mental Health Services of the Capital Region of Denmark, and Aarhus University Hospital - Psychiatry, Central Denmark Region. The funding sources had no role in the design and conduct of the study, the collection, management, analysis, and interpretation of the data, the preparation, review, or approval of the manuscript, or the decision to submit the manuscript for publication.

## Author Contribution

MMH and KSM (corresponding author) had full access to all the data in the study and take responsibility for the integrity of the data and the accuracy of the data analysis. MN, AAET, OM, LØ, and HRS designed the VIA study and applied for funding. KSM and MMH conceptualized the current study. JMB, MFK, MG, NH, AS, CBK, AKA, and LV recruited participants for the study and did the assessments of clinical symptoms. AAET supervised the clinical diagnosis, including the clinical diagnostic conferences. HS, TEL, and LØ were responsible for MR scanning. LKJ, KML, CBK, AKA, and LV scanned the participants. EH-T, WFCB, VI, MMH, and KSM analyzed the MR data. MMH manually edited the hippocampus under the supervision of KSM. EH-T, MMH, and KSM created the figures. MMH and KSM drafted the manuscript. All authors read and commented on the manuscript.

## Conflicts of interest disclosures

Hartwig R. Siebner has received honoraria as editor (Neuroimage Clinical) from Elsevier Publishers, Amsterdam, The Netherlands. He has received royalties as a book editor from Springer Publishers, Stuttgart, Germany; Oxford University Press, Oxford, UK; and Gyldendal Publishers, Copenhagen, Denmark. All disclosures are independent of this study.

## Notes

### Author Declarations

The National Committee on Health Research Ethics (Protocol number: H-16043682) and the Danish Data Protection Agency (ID: RHP-2017-003, I-suite: 05333) gave ethical approval for this work

