## Supplementary Material for "Children at familial high risk for schizophrenia or bipolar disorder show sex-specific differences in subcortical grey matter volumes"

#### Table of contents

##### Supplementary methods

Image acquisition

##### Supplementary Results

Dropout analyses

Whole sample pairwise group analyses

Role of lifetime Axis-I diagnosis in thalamus and amygdala volume differences

##### Supplementary Figures

**sFigure 1.** Flowchart of the inclusion and exclusion procedures

**sFigure 2.** Violin plots displaying subcortical volumes for each group for females and males

##### Supplementary Tables

**sTable 1.** Number of included participants of each structure after quality control of segmentation

**sTable 2**. Results from the drop-out analysis comparing VIA11 variables at age 11 years

**sTable 3.** Results from the drop-out analysis comparing VIA7 variables at age 7 years

**sTable 4.** Results from the subcortical volume analysis, additionally controlling for pubertal stage

**sTable 5.** Results of the pairwise analyses of subcortical volumes in males before and after TBV adjustment

**sTable 6.** Results of the pairwise analyses of subcortical volumes in females before and after TBV adjustment

**sTable 7.** Results of the pairwise analyses of subcortical volumes in the whole group before and after TBV adjustment

**sTable 8.** Results from the pairwise analysis of subcortical volumes with significant group-by-sex interactions, additionally controlling for lifetime Axis-I diagnosis

**sTable 9.** Results from the pairwise analysis comparing children with and without a lifetime Axis-I diagnosis

**sTable 10.** Results of the follow-up analysis of the unilateral volumes

**sTable 11.** Results of the analysis of group and group-by-sex differences in the R1-values

### Supplementary methods

#### Image acquisition

At DRCMR, images were acquired using a 3.0 Tesla Siemens Magnetom Prisma scanner equipped with a 64-channel head coil. The MP2RAGE was obtained with following parameters: TR = 6500 ms, TE = 3.49 ms, TI1 = 700 ms; TI2 = 2800 ms; flip angle 1 = 4 degrees; flip angle 2 = 6 degrees; field of view (FOV) = 260 x 260, 192 sagittal slices, 0.9 x 0.9 x 0.9 $mm^3$ voxels, acquisition time = 12:39. At CFIN, images were acquired using a 3.0 Tesla Siemens Magnetom Skyra with a 32-channel head coil. All sequence parameters were the same as for the Prisma, except for a TE=3.46 ms.

### Supplementary Results

#### Dropout analyses

Dropout analyses comparing the sample included in the present study with the full VIA7 cohort showed that included FHR-SZ children had higher CGAS scores (p<0.001) and lower total CBCL scores (p=0.041) than non-included FHR-SZ children. In addition, primary caregivers of included FHR-SZ children had higher PSP scores (p=0.013). Together, these findings indicate better functioning among included FHR-SZ children and their primary caregivers (Table S3).

#### Whole sample pairwise group analyses

For comparison with previous studies, we conducted post hoc whole-sample pairwise group analyses of the main effect of group, excluding the group-by-sex interaction (Table S7). Children at FHR-SZ had smaller hippocampal and accumbens volumes than PBC children (p≤0.040) and smaller amygdala, hippocampus, thalamus, accumbens, caudate, and pallidum volumes than FHR-BP children (p≤0.034). Children at FHR-BP had larger caudate volumes than PBC and FHR-SZ children (p≤0.016). In addition, relative thalamic and accumbens volumes were smaller in FHR-SZ children than in FHR-BP children (p≤0.040). Overall, the whole-sample findings largely reflected the sex-specific group differences reported in the main analyses

#### Role of lifetime Axis-I diagnosis in thalamus and amygdala volume differences

We examined whether the observed absolute thalamus and relative amygdala differences were related to lifetime Axis-I diagnosis by including diagnosis as a covariate in the models. The pattern of results remained unchanged. Males at FHR-SZ continued to show smaller thalamus volumes than both PBC and FHR-BP males (p≤0.006). In addition, females at FHR-BP had larger thalamus volumes than PBC females (p=0.017), and FHR-BP males showed larger relative amygdala volumes than PBC males (p=0.028). These findings indicate that the reported volume differences were not explained by Axis-I diagnosis (Table S8).

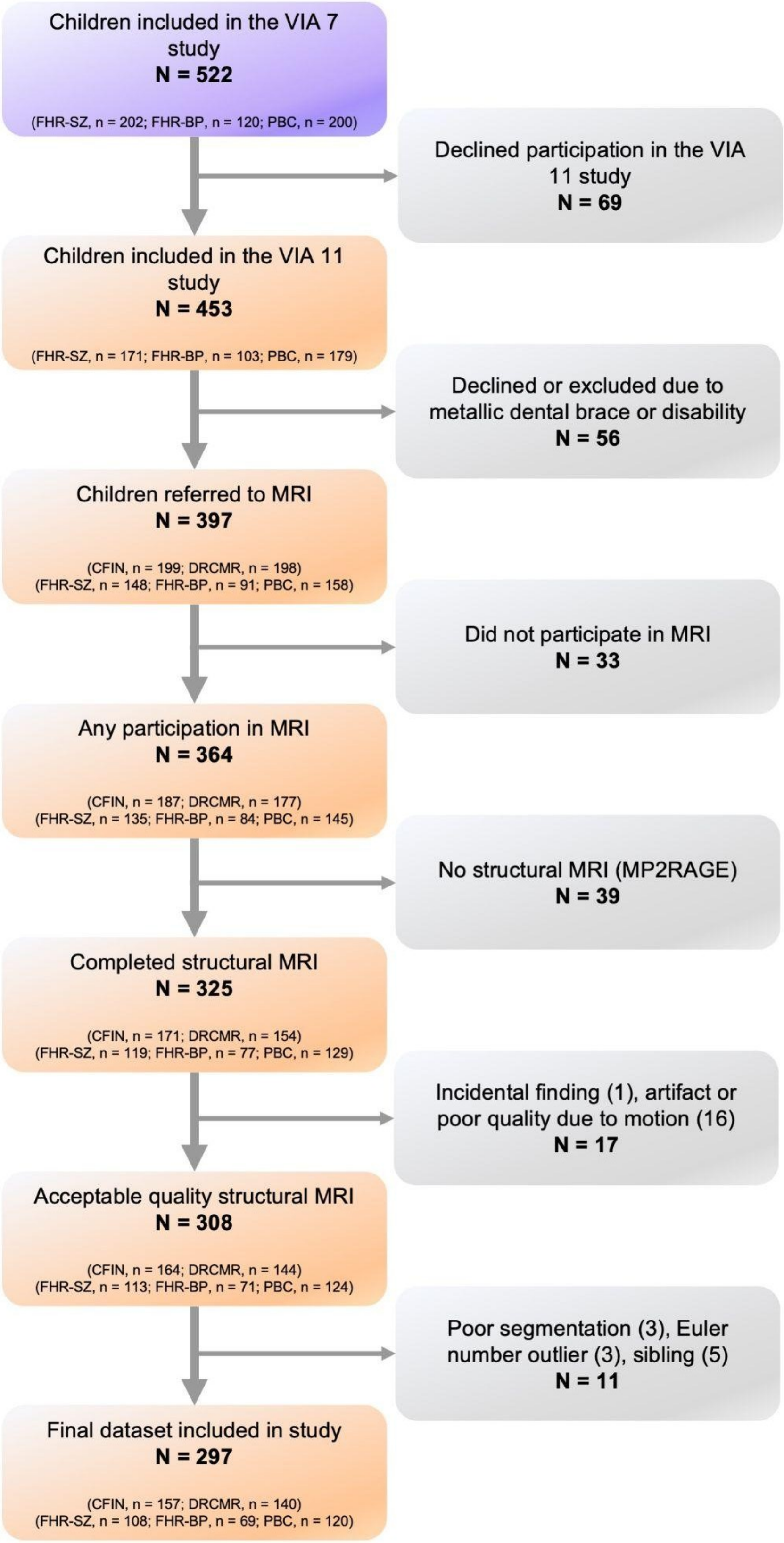

**Supplementary Figure 1.** Flowchart of the inclusion and exclusion procedures in the MRI study and in the current subcortical volume study. Abbreviations: CFIN: Center of Functionally Integrative Neuroscience, Aarhus. DRCMR: Danish Research Center for Magnetic Resonance. FHR-SZ: Familial high risk of schizophrenia. FHR-BP: Familial high risk of bipolar diagnosis. PBC: Population-based controls.

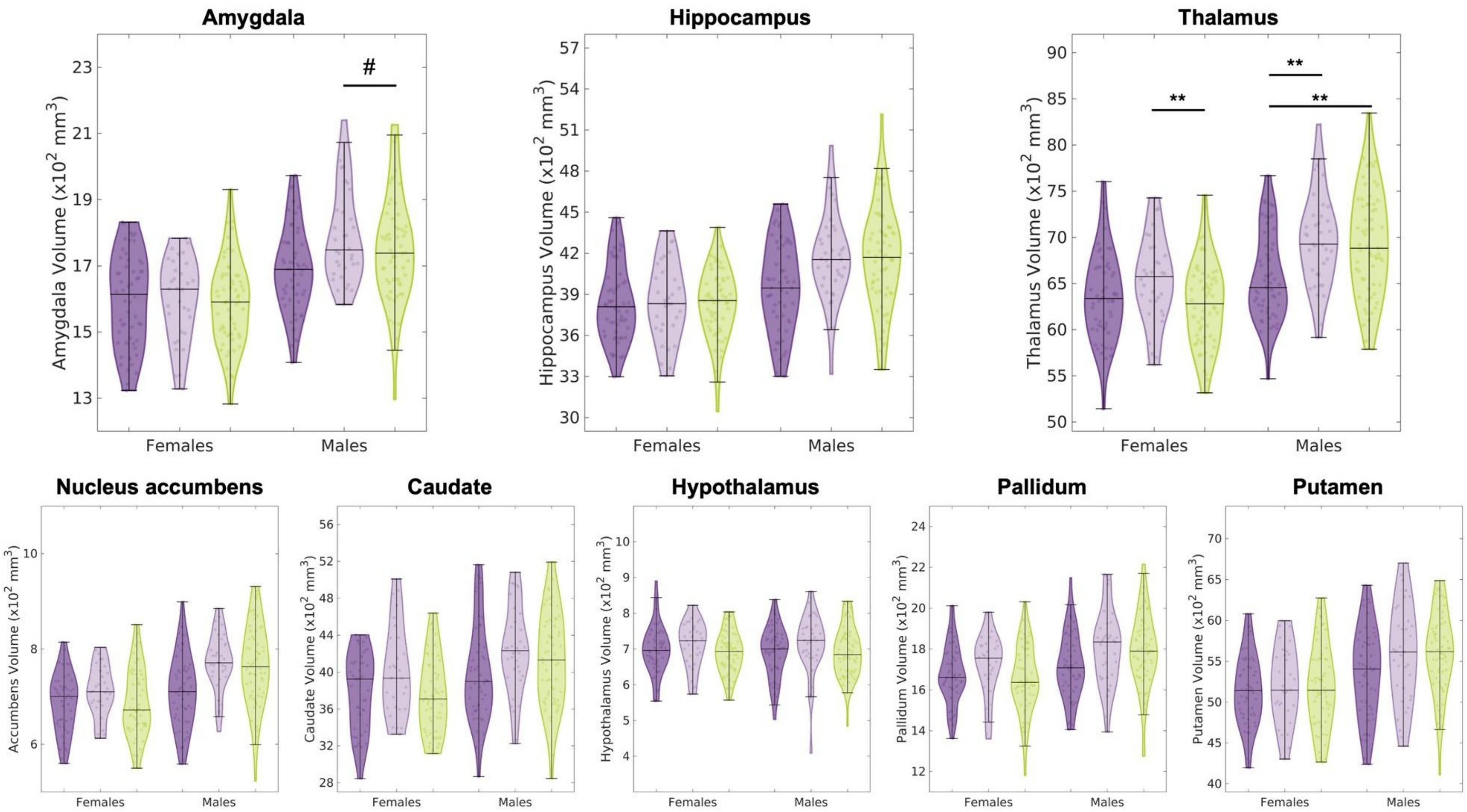

**Supplementary Figure 2.** Violin plots displaying bilateral amygdala, hippocampus, and thalamus (top row) and nucleus accumbens, caudate, hypothalamus, pallidum, and putamen volumes for females and males. The familial high risk for schizophrenia (FHR-SZ) or bipolar diagnosis (FHR-BP) groups are shown in, respectively, dark and light purple, while the population-based controls are shown in green. The horizontal lines indicate the median and the data range (excluding outliers). The dots within the violin plots show the individual data points. Statistical pairwise group differences are indicated at the .05 (*; # if the relative volume was significant) and .01 (**) uncorrected significance levels.

**Supplementary Table 1. Number of participants included for each structure (unilateral and bilateral) after quality control of segmentation.**

| | Total | FHR-SZ | FHR-BP | PBC | |
|---|---|---|---|---|---|
| Structures, N | (N=297) | (N=108) | (N=69) | (N=120) | *p* |
| **Amygdala** | | | | | |
| *Right* | 291 | 106 | 67 | 118 | 0.835 |
| *Left* | 293 | 107 | 67 | 119 | 0.442 |
| *Bilateral* | 290 | 106 | 66 | 118 | 0.768 |
| **Hippocampus** | | | | | |
| *Right* | 294 | 107 | 68 | 119 | 0.915 |
| *Left* | 293 | 108 | 68 | 117 | 0.262 |
| *Bilateral* | 291 | 107 | 68 | 116 | 0.146 |
| **Thalamus** | | | | | |
| *Right* | 296 | 108 | 69 | 119 | 0.477 |
| *Left* | 296 | 108 | 69 | 119 | 0.477 |
| *Bilateral* | 296 | 108 | 69 | 119 | 0.477 |
| **Accumbens** | | | | | |
| *Right* | 289 | 102 | 68 | 119 | 0.068 |
| *Left* | 284 | 101 | 66 | 117 | 0.341 |
| *Bilateral* | 277 | 96 | 65 | 116 | 0.175 |
| **Caudate** | | | | | |
| *Right* | 291 | 105 | 67 | 119 | 0.488 |
| *Left* | 270 | 93 | 64 | 113 | 0.089 |
| *Bilateral* | 269 | 93 | 63 | 113 | 0.344 |
| **Hypothalamus** | | | | | |
| *Bilateral* | 289 | 108 | 67 | 114 | 0.066 |
| **Pallidum** | | | | | |
| *Right* | 291 | 104 | 67 | 120 | 0.117 |
| *Left* | 282 | 101 | 65 | 116 | 0.528 |
| *Bilateral* | 279 | 98 | 65 | 116 | 0.112 |
| **Putamen** | | | | | |
| *Right* | 293 | 106 | 68 | 119 | 0.798 |
| *Left* | 291 | 106 | 66 | 119 | 0.252 |
| *Bilateral* | 288 | 104 | 66 | 118 | 0.375 |

Exclusion of one of the unilateral structures led to the exclusion of the structure bilaterally. P-values represent $\chi^2$ tests comparing the proportion of participants with available structural data relative to the original sample in each group. Abbreviations: FHR-SZ: familial high risk of schizophrenia, FHR-BP: familial high risk of bipolar disorder, PBC: population-based controls.

**Supplementary Table 2. Results from the drop-out analysis comparing VIA11 variables at age 11 years from participants included vs. not included in the current MRI study for each group.**

| | FHR-SZ | | | FHR-BP | | | PBC | | |
|---|---|---|---|---|---|---|---|---|---|
| **VIA11** | Not included | Included | p | Not included | Included | p | Not included | Included | p |
| Children, N | 63 | 108 | | 34 | 69 | | 59 | 120 | |
| Females, N (%) | 27 (42.9%) | 55 (50.9%) | 0.308 | 13 (38.2%) | 32 (46.4%) | 0.433 | 24 (40.7%) | 59 (49.2%) | 0.284 |
| CBCL Total, mean (SD)* | 27.9 (24.3) | 20.8 (17.6) | **0.016** | 23.9 (22.2) | 20.1 (20.7) | 0.320 | 13.9 (15.8) | 11.9 (10.5) | 0.500 |
| CGAS, mean (SD)* | 59.9 (15.6) | 67.4 (15.0) | **0.001** | 64.0 (15.0) | 70.2 (14.7) | **0.047** | 74.0 (14.0) | 75.7 (14.0) | 0.465 |
| Any Axis-I diagnosis, N (%)* | 38 (60.3%) | 54 (50.0%) | 0.237 | 21 (61.8%) | 33 (47.8%) | 0.183 | 18 (30.5%) | 32 (26.7%) | 0.540 |
| Primary caregiver's PSP, mean (SD)* | 66.6 (17.0) | 72.3 (16.5) | **0.013** | 71.9 (15.3) | 72.0 (15.7) | 0.972 | 84.5 (7.71) | 82.8 (11.2) | 0.463 |

Drop-out analysis tested differences in the VIA11 between participants included vs. non-included in the present study for each group (FHR-SZ, FHR-BP, PBC). Two-tailed t-tests were used to assess CBCL, CGAS, and PSP. A chi-square test assessed sex and prevalence of lifetime Axis-I diagnosis. Significant p-values are shown in bold. Abbreviations: FHR-SZ: Familial high risk for schizophrenia, FHR-BP: Familial high risk for bipolar diagnosis, PBC: Population-based controls, CBCL: Child Behavior Check List, CGAS: Children’s Global Assessment Scale, PSP: Personal and Social Performance. *Missing data in VIA11: CBCL (N=18), CGAS (N=4), Axis-I diagnosis (N=6), PSP (N=7).

**Supplementary Table 3. Results from the drop-out analysis comparing VIA7 variables at age 7 years from participants included vs. not included in the current MRI study for each group.**

| | FHR-SZ | | | FHR-BP | | | PBC | | |
|---|---|---|---|---|---|---|---|---|---|
| **VIA7** | Not included | Included | p | Not included | Included | p | Not included | Included | p |
| Children, N | 94 | 108 | | 51 | 69 | | 80 | 120 | |
| Females, N (%) | 38 (40.4%) | 55 (50.9%) | 0.135 | 24 (47.1%) | 32 (46.4%) | 0.941 | 34 (42.5%) | 59 (49.6%) | 0.354 |
| CBCL Total, mean (SD)* | 28.3 (23.3) | 26.3 (19.0) | 0.452 | 23.9 (20.6) | 23.0 (19.2) | 0.801 | 17.9 (15.8) | 16.4 (14.0) | 0.598 |
| CGAS, mean (SD)* | 64.4 (15.1) | 71.3 (15.0) | **<0.001** | 71.0 (15.7) | 75.4 (14.1) | 0.101 | 77.2 (14.0) | 78.0 (13.2) | 0.691 |
| Any Axis-I diagnosis, N (%)* | 43 (45.7%) | 34 (31.5%) | **0.041** | 21 (41.2%) | 21 (30.4%) | 0.165 | 12 (15.0%) | 18 (15.0%) | 0.911 |
| Primary caregiver's PSP, mean (SD)* | 70.5 (12.3) | 75.3 (15.2) | **0.007** | 71.9 (14.3) | 76.3 (13.9) | 0.056 | 84.0 (9.49) | 84.6 (8.92) | 0.740 |

Drop-out analysis tested differences in the VIA7 data between participants included vs. non-included in the present study for each group (FHR-SZ, FHR-BP, PBC). Two-tailed t-tests were used to assess CBCL, CGAS, and PSP. Chi-square tests assessed sex and prevalence of lifetime Axis-I diagnosis. Significant p-values are shown in bold. Abbreviations: FHR-SZ: Familial high risk for schizophrenia, FHR-BP: Familial high risk for bipolar diagnosis, PBC: Population-based controls, CBCL: Child Behavior Check List, CGAS: Children’s Global Assessment Scale, PSP: Personal and Social Performance. *Missing data: CBCL (N=28), CGAS (N=8), Axis-I diagnosis (N=8), PSP (N=10).

**Supplementary Table 4. Results from the subcortical volume analysis, additionally controlling for pubertal stage.**

| | Group | | Group-by-sex | | Age | | Sex | | MR-site | | Euler Number | | Pubertal stage | | Time Interval Tanner - MRI | |
|---|---|---|---|---|---|---|---|---|---|---|---|---|---|---|---|---|
| | *F* | *p* | *F* | *p* | *F* | *p* | *F* | *p* | *F* | *p* | *F* | *p* | *F* | *p* | *F* | *p* |
| **Without TBV** | | | | | | | | | | | | | | | | |
| Amygdala | 2.637 | 0.073 | 2.463 | 0.087 | 1.167 | 0.281 | 83.091 | <0.001 | 3.933 | 0.048 | 8.025 | 0.005 | 1.244 | 0.266 | 0.393 | 0.531 |
| Thalamus | 2.852 | 0.059 | 3.815 | **0.023** | 0.864 | 0.353 | 17.110 | <0.001 | 2.330 | 0.128 | 26.528 | <0.001 | 0.583 | 0.446 | 0.955 | 0.329 |
| **With TBV** | | | | | | | | | | | | | | | | |
| Amygdala | 1.241 | 0.291 | 4.193 | **0.016** | 3.795 | 0.052 | 17.583 | <0.001 | 0.595 | 0.441 | 0.091 | 0.764 | 0.121 | 0.728 | 0.437 | 0.509 |
| Thalamus | 1.854 | 0.159 | 0.244 | 0.784 | 1.771 | 0.184 | 9.153 | 0.003 | 0.004 | 0.948 | 15.542 | <0.001 | 0.206 | 0.650 | 0.780 | 0.378 |

Each row represents a separate ANCOVA model, examining group-by-sex differences. All models included the covariates: age, sex, MR-site, Euler number, pubertal stage, and time interval between Tanner stage assessment and MRI. Significant group-by-sex effects at the uncorrected 0.05 level are shown in bold. Number of male/female children in each group: FHR-SZ: n=53/55, FHR-BP: n=37/32, PBC: n=61/59. Abbreviations: TBV: total brain volume.

#### Supplementary Table 5. Results of the pairwise analyses of subcortical volumes in males before and after TBV adjustment.

| | Least square mean volumes | | | FHR-SZ vs. PBC | | | | FHR-BP vs. PBC | | | | FHR-SZ vs. FHR-BP | | | |
|---|---|---|---|---|---|---|---|---|---|---|---|---|---|---|---|
| | FHR-SZ | FHR-BP | PBC | *t* | *p* | *d* | $BF_{10}$ | *t* | *p* | *d* | $BF_{10}$ | *t* | *p* | *d* | $BF_{10}$ |
| **Without TBV** | | | | | | | | | | | | | | | |
| Amygdala | 1693.8 | 1794.0 | 1738.1 | -1.610 | 0.110 | -0.311 | 0.728 | 1.783 | 0.077 | 0.392 | 1.008 | -3.100 | **0.002** | -0.703 | 15.647 |
| Hippocampus | 3970.8 | 4159.5 | 4138.1 | -2.619 | **0.010** | -0.504 | 4.580 | 0.301 | 0.764 | 0.065 | 0.229 | -2.585 | **0.011** | -0.569 | 4.600 |
| Thalamus* | 6602.6 | 6949.7 | 6908.9 | -3.270 | **0.001** | -0.626 | 25.397 | 0.390 | 0.697 | 0.083 | 0.267 | -3.225 | **0.002** | -0.709 | 23.826 |
| Accumbens | 715.5 | 766.0 | 766.1 | -3.495 | **0.001** | -0.685 | 29.278 | -0.006 | 0.995 | -0.001 | 0.232 | -2.983 | **0.003** | -0.684 | 21.412 |
| Caudate | 4018.4 | 4203.7 | 4115.4 | -1.008 | 0.315 | -0.197 | 0.322 | 0.815 | 0.416 | 0.180 | 0.407 | -1.639 | 0.103 | -0.377 | 0.614 |
| Hypothalamus | 691.3 | 713.7 | 697.2 | -0.433 | 0.666 | -0.085 | 0.222 | 1.093 | 0.276 | 0.238 | 0.435 | -1.453 | 0.148 | -0.322 | 0.577 |
| Pallidum | 1731.1 | 1788.6 | 1780.3 | -1.506 | 0.134 | -0.297 | 0.816 | 0.228 | 0.820 | 0.050 | 0.242 | -1.521 | 0.130 | -0.348 | 0.695 |
| Putamen | 5395.4 | 5555.2 | 5581.2 | -1.809 | 0.073 | -0.355 | 1.487 | -0.228 | 0.820 | -0.050 | 0.231 | -1.357 | 0.177 | -0.305 | 0.398 |
| **With TBV** | | | | | | | | | | | | | | | |
| Amygdala* | 1712.9 | 1772.3 | 1740.2 | 1.405 | 0.162 | 0.283 | 0.478 | 2.799 | **0.006** | 0.616 | 11.349 | -1.426 | 0.156 | -0.333 | 0.588 |
| Hippocampus | 4092.0 | 4115.3 | 4060.2 | -0.587 | 0.558 | -0.118 | 0.229 | 0.404 | 0.687 | 0.086 | 0.242 | -0.900 | 0.370 | -0.205 | 0.448 |
| Thalamus | 6819.6 | 6866.7 | 6774.5 | -0.699 | 0.485 | -0.140 | 0.283 | 0.684 | 0.495 | 0.146 | 0.335 | -1.259 | 0.210 | -0.286 | 0.434 |
| Accumbens | 755.6 | 758.9 | 734.5 | -1.716 | 0.088 | -0.351 | 0.590 | 0.251 | 0.802 | 0.055 | 0.236 | -1.729 | 0.086 | -0.406 | 1.436 |
| Caudate | 4054.3 | 4149.0 | 4129.0 | 0.958 | 0.340 | 0.196 | 0.306 | 0.974 | 0.332 | 0.215 | 0.476 | -0.080 | 0.936 | -0.019 | 0.252 |
| Hypothalamus | 693.1 | 708.2 | 700.3 | 0.533 | 0.595 | 0.108 | 0.236 | 1.042 | 0.299 | 0.227 | 0.437 | -0.518 | 0.605 | -0.119 | 0.408 |
| Pallidum | 1754.8 | 1767.3 | 1779.2 | 0.884 | 0.378 | 0.183 | 0.327 | 0.427 | 0.670 | 0.094 | 0.242 | 0.378 | 0.706 | 0.089 | 0.256 |
| Putamen | 5513.8 | 5489.1 | 5528.8 | 0.164 | 0.870 | 0.034 | 0.220 | -0.252 | 0.801 | -0.055 | 0.241 | 0.381 | 0.704 | 0.089 | 0.259 |

Pairwise t-tests of least square mean volumes ($mm^3$) of subcortical grey matter structures for *males* in the three groups. *Significant group-by-sex effect in the a priori ANCOVA (q<0.05). Significant pairwise effects at the uncorrected (p-values) 0.05 level are shown in bold. Abbreviations: FHR-SZ: familial high risk of schizophrenia; FHR-BP: familial high risk of bipolar disorder; PBC: population-based controls; *d:* Cohen's *d*; $BF_{10}$: Bayes Factor, TBV: total brain volume.

#### Supplementary Table 6. Results of the pairwise analyses of subcortical volumes in females before and after TBV adjustment.

| | Least square mean volumes | | | FHR-SZ vs. PBC | | | | FHR-BP vs. PBC | | | | FHR-SZ vs. FHR-BP | | | |
|---|---|---|---|---|---|---|---|---|---|---|---|---|---|---|---|
| | FHR-SZ | FHR-BP | PBC | *t* | *p* | *d* | $BF_{10}$ | *t* | *p* | *d* | $BF_{10}$ | *t* | *p* | *d* | $BF_{10}$ |
| **Without TBV** | | | | | | | | | | | | | | | |
| Amygdala | 1595.6 | 1598.3 | 1588.2 | 0.291 | 0.772 | 0.055 | 0.206 | 0.343 | 0.732 | 0.076 | 0.232 | -0.093 | 0.926 | -0.021 | 0.242 |
| Hippocampus | 3834.6 | 3840.9 | 3829.7 | 0.094 | 0.925 | 0.018 | 0.198 | 0.184 | 0.854 | 0.041 | 0.235 | -0.104 | 0.918 | -0.023 | 0.237 |
| Thalamus | 6357.0 | 6536.5 | 6280.3 | 0.888 | 0.376 | 0.168 | 0.264 | 2.523 | **0.013*** | 0.560 | 3.112 | -1.759 | 0.081 | -0.392 | 0.994 |
| Accumbens | 687.0 | 712.0 | 686.3 | 0.050 | 0.960 | 0.010 | 0.212 | 1.773 | 0.079 | 0.401 | 0.698 | -1.677 | 0.096 | -0.390 | 1.204 |
| Caudate | 3800.1 | 4011.6 | 3745.2 | 0.618 | 0.538 | 0.126 | 0.270 | 2.637 | **0.009** | 0.611 | 3.486 | -2.019 | **0.046** | -0.485 | 1.537 |
| Hypothalamus | 696.4 | 714.0 | 689.4 | 0.599 | 0.550 | 0.114 | 0.244 | 1.767 | 0.079 | 0.398 | 0.762 | -1.261 | 0.209 | -0.284 | 0.436 |
| Pallidum | 1676.4 | 1721.0 | 1655.2 | 0.685 | 0.494 | 0.135 | 0.263 | 1.853 | 0.066 | 0.420 | 0.890 | -1.211 | 0.228 | -0.284 | 0.502 |
| Putamen | 5149.1 | 5219.5 | 5176.3 | -0.300 | 0.765 | -0.057 | 0.204 | 0.406 | 0.686 | 0.091 | 0.246 | -0.650 | 0.517 | -0.148 | 0.311 |
| **With TBV** | | | | | | | | | | | | | | | |
| Amygdala | 1593.3 | 1573.2 | 1604.2 | -0.620 | 0.537 | -0.118 | 0.236 | -1.492 | 0.138 | -0.335 | 0.859 | 0.965 | 0.336 | 0.217 | 0.303 |
| Hippocampus | 3826.9 | 3791.9 | 3864.5 | -0.950 | 0.344 | -0.183 | 0.280 | -1.541 | 0.126 | -0.353 | 0.763 | 0.751 | 0.454 | 0.170 | 0.282 |
| Thalamus | 6347.9 | 6446.7 | 6339.3 | 0.161 | 0.872 | 0.030 | 0.203 | 1.686 | 0.094 | 0.379 | 0.846 | -1.556 | 0.122 | -0.348 | 0.718 |
| Accumbens | 687.2 | 700.2 | 692.7 | -0.529 | 0.598 | -0.106 | 0.246 | 0.627 | 0.532 | 0.144 | 0.288 | -1.068 | 0.288 | -0.250 | 0.560 |
| Caudate | 3794.7 | 3948.1 | 3784.0 | 0.136 | 0.892 | 0.028 | 0.213 | 1.812 | 0.072 | 0.428 | 0.963 | -1.654 | 0.101 | -0.400 | 0.894 |
| Hypothalamus | 695.7 | 705.5 | 694.7 | 0.100 | 0.921 | 0.019 | 0.207 | 0.854 | 0.394 | 0.196 | 0.376 | -0.779 | 0.437 | -0.177 | 0.293 |
| Pallidum | 1672.6 | 1697.7 | 1671.2 | 0.060 | 0.952 | 0.012 | 0.206 | 0.945 | 0.346 | 0.217 | 0.352 | -0.870 | 0.386 | -0.205 | 0.379 |
| Putamen | 5146.9 | 5157.2 | 5211.3 | -0.808 | 0.420 | -0.154 | 0.246 | -0.572 | 0.568 | -0.129 | 0.266 | -0.108 | 0.914 | -0.025 | 0.244 |

Pairwise t-tests of least square mean volumes ($mm^3$) of subcortical grey matter structures for fe*males* in the three groups. *Significant group-by-sex effect in the a priori ANCOVA (q<0.05). Significant pairwise effects at the uncorrected (p-values) 0.05 level are shown in bold. Abbreviations: FHR-SZ: familial high risk of schizophrenia; FHR-BP: familial high risk of bipolar disorder; PBC: population-based controls; *d:* Cohen's *d*; $BF_{10}$: Bayes Factor, TBV: total brain volume.

#### Supplementary Table 7. Results of the pairwise analyses of subcortical volumes in the whole group before and after TBV adjustment.

| | **Least square mean volumes** | | | **FHR-SZ vs. PBC** | | | **FHR-BP vs. PBC** | | | **FHR-SZ vs. FHR-BP** | | |
|---|---|---|---|---|---|---|---|---|---|---|---|---|
| | FHR-SZ | FHR-BP | PBC | t | p | d | t | p | d | t | p | d |
| **Without TBV** | | | | | | | | | | | | |
| Amygdala | 1645.5 | 1694.1 | 1665.6 | -1.070 | 0.286 | -0.144 | 1.323 | 0.187 | 0.204 | -2.213 | **0.028** | -0.349 |
| Hippocampus | 3901.4 | 4003.2 | 3986.5 | -2.062 | **0.040** | -0.279 | 0.354 | 0.723 | 0.055 | -2.136 | **0.034** | -0.331 |
| Thalamus | 6479.8 | 6740.0 | 6597.7 | -1.841 | 0.067 | -0.247 | 1.956 | 0.051 | 0.297 | -3.511 | **0.001** | -0.554 |
| Accumbens | 699.9 | 739.5 | 725.6 | -2.634 | **0.009** | -0.367 | 1.268 | 0.206 | 0.076 | -3.495 | **0.001** | -0.565 |
| Caudate | 3898.8 | 4113.1 | 3935.0 | -0.552 | 0.581 | -0.078 | 2.415 | **0.016** | 0.382 | -2.800 | **0.006** | -0.459 |
| Hypothalamus | 694.2 | 711.9 | 693.4 | 0.082 | 0.934 | 0.011 | 1.823 | 0.069 | 0.282 | -1.730 | 0.085 | -0.270 |
| Pallidum | 1698.8 | 1757.3 | 1719.9 | -0.938 | 0.349 | -0.130 | 1.475 | 0.141 | 0.230 | -2.238 | **0.026** | -0.360 |
| Putamen | 5263.6 | 5395.7 | 5382.8 | -1.748 | 0.082 | -0.237 | 0.165 | 0.869 | 0.026 | -1.663 | 0.097 | -0.263 |
| **With TBV** | | | | | | | | | | | | |
| Amygdala | 1666.0 | 1672.3 | 1661.7 | 0.333 | 0.739 | 0.045 | 0.716 | 0.474 | 0.111 | -0.412 | 0.681 | -0.066 |
| Hippocampus | 3942.5 | 3961.8 | 3983.6 | -1.261 | 0.208 | -0.171 | -0.589 | 0.556 | -0.091 | -0.509 | 0.611 | -0.081 |
| Thalamus | 6562.6 | 6660.2 | 6584.4 | -0.538 | 0.591 | -0.073 | 1.651 | 0.100 | 0.252 | -2.065 | **0.040** | -0.324 |
| Accumbens | 710.4 | 731.0 | 724.6 | -1.792 | 0.074 | -0.251 | 0.725 | 0.469 | 0.113 | -2.225 | **0.027** | -0.365 |
| Caudate | 3960.8 | 4058.7 | 3934.8 | 0.461 | 0.645 | 0.065 | 1.957 | 0.052 | 0.310 | -1.471 | 0.142 | -0.245 |
| Hypothalamus | 699.1 | 705.4 | 693.4 | 0.686 | 0.494 | 0.093 | 1.271 | 0.205 | 0.197 | -0.655 | 0.513 | -0.104 |
| Pallidum | 1722.9 | 1737.7 | 1716.2 | 0.372 | 0.710 | 0.052 | 1.073 | 0.284 | 0.168 | -0.710 | 0.478 | -0.116 |
| Putamen | 5323.4 | 5334.7 | 5371.1 | -0.803 | 0.422 | -0.110 | -0.542 | 0.589 | -0.084 | -0.161 | 0.872 | -0.026 |

Pairwise t-tests of least square mean volumes ($mm^3$) of subcortical grey matter structures in the three groups. Significant pairwise effects at the uncorrected (p-values) 0.05 level are shown in bold. Abbreviations: FHR-SZ: familial high risk of schizophrenia; FHR-BP: familial high risk of bipolar disorder; PBC: population-based controls; *d:* Cohen's *d*; $BF_{10}$: Bayes Factor, TBV: total brain volume.

**Supplementary Table 8. Results from the pairwise analysis of subcortical volumes with significant group-by-sex interactions, additionally controlling for lifetime Axis-I diagnosis.**

| | Least square mean volumes (mm$^3$) | | | FHR-SZ vs. PBC | | | FHR-BP vs. PBC | | | FHR-SZ vs. FHR-BP | | |
|---|---|---|---|---|---|---|---|---|---|---|---|---|
| | FHR-SZ | FHR-BP | PBC | *d* | *t* | *p* | *d* | *t* | *p* | *d* | *t* | *p* |
| **Females** | | | | | | | | | | | | |
| Thalamus volume | 6359 | 6554 | 6308 | 0.113 | 0.578 | 0.564 | 0.540 | 2.427 | **0.017** | -0.426 | -1.882 | 0.062 |
| Amygdala volume† | 1594 | 1574 | 1605 | -0.114 | -0.580 | 0.563 | -0.333 | -1.484 | 0.140 | 0.220 | 0.961 | 0.338 |
| **Males** | | | | | | | | | | | | |
| Thalamus volume | 6602 | 6963 | 6872 | -0.551 | -2.775 | **0.006** | 0.185 | 0.829 | 0.409 | -0.735 | -3.316 | **0.001** |
| Amygdala volume† | 1743 | 1768 | 1719 | -0.246 | -1.196 | 0.234 | 0.511 | 2.225 | **0.028** | 0.265 | 1.124 | 0.263 |

Follow-up pairwise analysis using t-tests on the estimated marginal means obtained from lsmeans comparing the FHR-SZ vs. PBC, FHR-BP vs. PBC, and FHR-SZ vs. FHR-BP groups in females and males separately. Lsmeans were obtained from ANCOVA models of group differences in brain measures, controlling for age, sex, site, Euler number, and lifetime Axis-I diagnosis. Significant group differences at the uncorrected 0.05 level are shown in bold. Abbreviations: FHR-SZ: familial high risk for schizophrenia; FHR-BP: familial high risk for bipolar disorder; PBC: population-based controls, *d*: Cohen's *d*. Missing data: Axis-I diagnosis: PBC males: n=2, FHR-SZ male: n=1, FHR-SZ female: n=1). †*Controlling for total brain volume.*

**Supplementary Table 9. Results from the pairwise analysis comparing children with (+) and without (-) a lifetime Axis-I diagnosis within each group.**

| | Least square mean volumes (mm$^3$) | | | | | | FHR-SZ+ vs. FHR-SZ- | | FHR-BP+ vs. FHR-BP- | | PBC+ vs. PBC- | |
|---|---|---|---|---|---|---|---|---|---|---|---|---|
| | FHR-SZ+ | FHR-SZ- | FHR-BP+ | FHR-BP- | PBC+ | PBC- | *t* | *p* | *t* | *p* | *t* | *p* |
| **Females** | | | | | | | | | | | | |
| Thalamus volume | 6454 | 6271 | 6572 | 6521 | 6289 | 6276 | 1.457 | 0.152 | 0.278 | 0.783 | 0.093 | 0.926 |
| Amygdala volume† | 1583 | 1612 | 1609 | 1592 | 1600 | 1585 | -0.942 | 0.351 | 0.583 | 0.565 | 0.550 | 0.584 |
| **Males** | | | | | | | | | | | | |
| Thalamus volume | 6485 | 6660 | 6889 | 7070 | 6886 | 6929 | -1.426 | 0.161 | -1.018 | 0.317 | -0.263 | 0.794 |
| Amygdala volume† | 1709 | 1680 | 1788 | 1799 | 1791 | 1725 | 1.119 | 0.269 | -0.230 | 0.820 | 2.501 | **0.015** |

Follow-up pairwise group analysis comparing the FHR-SZ+ vs. FHR-SZ-, FHR-BP+ vs. FHR-BP-, and PBC+ vs. PBC- groups in females and males separately. Significant group differences at the uncorrected 0.05 level are shown in bold. Number of male/female children in each group: FHR-SZ+: n=25/29, FHR-SZ-: n=27/25, FHR-BP+: n=22/11, FHR-BP-: n=15/21, PBC+: n=16/16, PBC-: n=43/44. Abbreviations: FHR-SZ: familial high risk for schizophrenia; FHR-BP: familial high risk for bipolar disorder; PBC: population-based controls. Missing data: Axis-I diagnosis: PBC males: n=2, FHR-SZ male: n=1, FHR-SZ female: n=1). †*Controlling for total brain volume.*

#### Supplementary Table 10. Results of the follow-up analysis of the unilateral volumes.

| | Least square mean volumes (mm³) | | | FHR-SZ vs. PBC | | | FHR-BP vs. PBC | | | FHR-SZ vs. FHR-BP | | |
|---|---|---|---|---|---|---|---|---|---|---|---|---|
| | FHR-SZ | FHR-BP | PBC | *t* | *p* | *d* | *t* | *p* | *d* | *t* | *p* | *d* |
| **Females** | | | | | | | | | | | | |
| Right thalamus | 6269.8 | 6439.0 | 6190.2 | 0.947 | 0.345 | 0.179 | 2.517 | **0.013** | 0.559 | -1.703 | 0.091 | -0.380 |
| Left thalamus | 6444.2 | 6634.1 | 6370.5 | 0.822 | 0.412 | 0.155 | 2.500 | **0.014** | 0.555 | -1.791 | 0.076 | -0.399 |
| **Males** | | | | | | | | | | | | |
| Right thalamus | 6513.4 | 6847.3 | 6820.9 | -3.385 | **0.001** | -0.648 | 0.260 | 0.795 | 0.056 | -3.199 | **0.002** | -0.703 |
| Left thalamus | 6691.8 | 7052.1 | 6997.0 | -3.116 | **0.002** | -0.596 | 0.505 | 0.614 | 0.108 | -3.203 | **0.002** | -0.704 |
| Right amygdala, adjusted for TBV | 1791.3 | 1826.9 | 1775.0 | 1.360 | 0.176 | 0.274 | 2.296 | **0.023** | 0.499 | -0.975 | 0.331 | -0.225 |
| Left amygdala, adjusted for TBV | 1672.8 | 1710.1 | 1649.3 | 1.148 | 0.253 | 0.231 | 2.730 | **0.007** | 0.596 | -1.583 | 0.116 | -0.365 |

Pairwise comparison of unilateral measures of the structures that were found to significantly differ between FHR and PBC children. Abbreviations: FHR-SZ: familial high risk of schizophrenia, FHR-BP: familial high risk of bipolar disorder, PBC: population-based controls, *d:* Cohen's *d*, TBV: total brain volume.

#### Supplementary Table 11. Results of the analysis of group and group-by-sex differences in the R1-values.

| | | Group | | | | Group*sex | | | |
|---|---|---|---|---|---|---|---|---|---|
| Structures | Model | *F* | *p* | *q* | $BF_{10}$ | *F* | *p* | *q* | $BF_{10}$ |
| Amygdala | 1 | 2.584 | 0.077 | - | - | 3.989 | **0.020** | 0.059 | 2.281 |
| Hippocampus | 1 | 1.678 | 0.189 | - | - | 3.148 | **0.048** | 0.072 | 0.942 |
| Thalamus | 1 | 0.024 | 0.976 | - | - | 1.872 | 0.156 | - | - |
| Thalamus | 2 | 1.294 | 0.270 | 0.270 | 0.132 | - | - | - | - |
| Accumbens | 1 | 3.226 | 0.041 | - | - | 4.067 | **0.018** | 0.069 | 2.242 |
| Caudate | 1 | 0.332 | 0.718 | - | - | 2.161 | 0.117 | - | - |
| Caudate | 2 | 0.598 | 0.551 | 0.642 | 0.073 | - | - | - | - |
| Pallidum | 1 | 0.220 | 0.803 | - | - | 1.259 | 0.286 | - | - |
| Pallidum | 2 | 0.817 | 0.443 | 0.620 | 0.093 | - | - | - | - |
| Putamen | 1 | 0.536 | 0.586 | - | - | 1.141 | 0.321 | - | - |
| Putamen | 2 | 0.339 | 0.713 | 0.713 | 0.057 | - | - | - | - |

Results of ANCOVA: Model 1: group-by-sex, group, sex, age, site, and Total Euler Number as covariates. Model 2: group, sex, age, site, and Total Euler Number as covariates. q: FDR-corrected q-values are reported for the group-by-sex model (Model 1) when $p < 0.05$; otherwise, they are reported for the group model (Model 2). Significant q values are shown in bold. Abbreviations: $BF_{10}$: Bayes Factor
